# Relationship Between Ratio of Low-Density Lipoprotein Cholesterol subtypes and Risk of Chronic Kidney Disease: The mediating role of inflammation

**DOI:** 10.1101/2025.03.17.25323166

**Authors:** Tingyu Wu, Rong Ma, Zhiqiang Duan, Zhuoxing Li, Ping Zhou, Wan Li, Qiyuan Liang, Chunlin Yu, Donglin Liu, Haiyan Yu, Guifei Deng, Yujun Tang, Xiang Xiao

## Abstract

**Objective:** This study aimed to investigate the relationship between the Small Dense Low-Density Lipoprotein Cholesterol/Large Buoyant Low-Density Lipoprotein Cholesterol Ratio (SLR) and the risk of Chronic Kidney Disease (CKD) in the U.S. population.

**Methods:** Data were recruited from the National Health and Nutrition Examination Survey (NHANES) database from 2009 to 2018. Restricted cubic spline (RCS) plots were used to assess the dose-effect relationship between SLR and the risk of CKD. Multiple logistic regression analysis was conducted to explore the relationship between SLR and the risk of CKD. Stratified analysis was performed to evaluate the consistency of the results. Mediation analysis explores the mediating roles of inflammatory indices Systemic Immune-Inflammation Index and Systemic Inflammation Response Index in SLR associated with the risk of CKD.

**Results:** A total of 11,905 participants were enrolled. The RCS showed an increased risk of CKD with higher SLR levels (nknot=5, Non-line *P* value <0.01). Multiple logistic regression analysis revealed that individuals in the T3 group had a 54% higher risk of CKD compared to the T1 group (OR = 1.54; 95% CI, 1.16, 2.06; *P* = 0.004). Additionally, per standard deviation (per-SD) increase in SLR, the risk of CKD increased by 16% (OR = 1.16; 95% CI, 1.05-1.29; *P* = 0.005). The relationship between SLR and the risk of CKD exerts a significant mediating effect through SII or SIRI.

**Conclusion:** In the general population, an elevated SLR is associated with a higher risk of CKD, and inflammatory plays a significant role in this process.

## 1. Introduction

Chronic Kidney Disease (CKD) has become one of the significant diseases affecting human health globally, with increasing incidence year by year, imposing a heavy burden on society and individuals(1–4). Exploring the pathogenesis of CKD and related risk factors is crucial for the early prevention, diagnosis, and treatment of the disease.

Among the various factors associated with the progression of CKD, abnormal lipid metabolism has been a major concern(5–7). Small dense Low-Density Lipoprotein cholesterol (sdLDL-C), as a special type of cholesterol, has metabolic and biological characteristics different from traditional LDL cholesterol (LDL-C)(8, 9). A study indicated a significant association between sdLDL-C and the prognosis of kidney and cardiovascular mortality in patients with diabetic nephropathy(10). Large buoyant Low-Density Lipoprotein cholesterol (lbLDL-C) is a subfraction of LDLc with smaller density and larger granules, and its causal role in the occurrence and development of diseases is not clear. However, it is certain that the physiologic roles of different subtypes of LDL-C are inconsistent. Additionally, research has demonstrated a close relationship between LDL-C and its subtypes and the occurrence of inflammatory. For instance, sdLDL-C was associated with low-grade inflammation and can induce inflammatory signaling pathways(11, 12). Inflammation plays a crucial role in the progression of CKD. Current evidence suggests that inflammation, regardless of its cause, can alter or disrupt renal microcirculation and perfusion distribution, leading to kidney injury and promoting the progression of CKD(13).

Abnormalities in lipid metabolism play a crucial role in the progression of CKD. For instance, hyperlipidemia has been established as the most common independent risk factor for CKD (14).Recent study has found a close relationship between the Small Dense Low-Density Lipoprotein Cholesterol/Large Buoyant Low-Density Lipoprotein Cholesterol Ratio (SLR),and the occurrence of diseases, suggesting heterogeneity among LDL-C subfractions (15–17) In healthy individuals and patients with metabolic syndrome (MetS), there is no significant difference in LDL-C levels, but there are differences in the ability to identify MetS among LDL subclasses (18). In HIV-infected patients, total cholesterol and LDL-C concentrations are similar, but patients with lipodystrophy, have higher sdLDL-C concentrations, and the SLR ratio plays an important role in identifying lipodystrophy(16). Therefore, due to the potential different roles of LDL-C subfractions in the diseases, SLR may be a better predictor of diseases.

Therefore, due to the potential different roles of LDL-C subfractions in the occurrence of diseases, SLR may be a better predictor of diseases. Clarifying the relationship between SLR levels and the risk of CKD may not only help to further understand the pathogenesis of CKD but also may provide new insights and evidence for the risk assessment of CKD.

Therefore, this study aims to analyze the potential relationship between SLR and the risk of CKD in the general population.

## 2. Methods

### 2.1. Study Design and Participants

The study utilized data from the National Health and Nutrition Examination Survey (NHANES) database for the period 2009 to 2018. NHANES focuses on gathering detailed demographic, health, and nutritional data through personal interviews and standardized physical exams at Mobile Examination Centers (MECs). The survey also evaluates the health and nutritional conditions of non-hospitalized individuals in the United States.

To ensure the study’s accuracy, specific diagnostic criteria were applied to identify the presence of CKD. These criteria align with the guidelines set forth by Kidney Disease Improving Global Outcomes (KDIGO) (19). These criteria included a urinary albumin-to-creatinine ratio (ACR) exceeding 30 mg/g or/and an estimated glomerular filtration rate (eGFR) below 60 ml/min/ 1.73m^2^. The exclusion criteria for this study were: 1) age < 20 years, 2) pregnancy, 3) dialysis, 4) missing CKD diagnostic data, 5) missing LDL-C and triglyceride (TG) data. According to literature (20), lbLDL-C was calculated using the equation lbLDL-C=1.43 × LDL-C-(0.14 ×(ln(TG)× LDL-C)- 8.99), and sdLDL-C was calculated using the equation sdLDL-C = LDL-C - lbLDL-C. Systemic Immune-Inflammation Index (SII) = Platelet Count × Neutrophil Count / Lymphocyte Count. Systemic Inflammation Response Index (SIRI) = (Neutrophil Count × Monocyte Count) / Lymphocyte Count. Definitions of smoking, alcohol, hypertension, hyperlipidemia, diabetes, anemia, etc., were provided in **Supplementary-table 1**.

The National Health and Nutrition Examination Survey (NHANES) is a research initiative carried out by the National Center for Health Statistics within the Centers for Disease Control and Prevention (CDC). It received approval from the Institutional Review Board of the National Center for Health Statistics. The study adheres to the ethical principles outlined in the 1964 Helsinki Declaration and its later revisions, having also obtained approval from the National Ethics Review Board for Health Statistics Research. Informed consent was provided by all participants (21).

### 2.2. Statistical Analysis

In accordance with the guidelines from the US Centers for Disease Control and Prevention, weighted samples were utilized in this study. A restricted cubic spline (RCS) plot was employed to assess the relationship between SLR and CKD risk in the general population. Participants were grouped into three categories based on SLR: lower (T1), medium (T2), and higher (T3), using a tertile approach. Weighted univariate logistic regression models analyzed the risk factors associated with CKD in the general population. Furthermore, weighted multivariable logistic regression models were applied to examine the linkage between SLR and CKD risk, with consistency of results confirmed through stratified analysis. Mediation analysis explores the mediating roles of inflammatory indices SII and SIRI in SLR associated with the risk of CKD. The weights were calculated by identifying the smallest variable subset in the study and applying the relevant weights, which were subsequently combined across different years. All statistical analyses were conducted using R software version 4.3.1, with a significance threshold set at two-sided *P < 0.05* for all tests.

## 3. Result

### 3.1 Baseline characteristics

In our study, we utilized data from 49,693 individuals registered in the NHANES database from 2009 to 2018. Among this dataset, we included 11,905 individuals **(Figure 1)**. The mean age of the study population was 48.29 years, with males accounting for 48.52%. Baseline characteristics were shown in the table **(Table 1)**. There were no significant differences in the proportions of alcohol use and anemia patients among the groups (*P* > 0.05). Significant differences were observed in age, albumin creatinine ratio (ACR), body mass index (BMI), estimated glomerular filtration rate (eGFR), gender, smoking, serum albumin, hyperlipidemia, hypertension, diabetes mellitus (DM), and renin-angiotensin system inhibitors (RAASi) proportions (*P* < 0.05). Individuals with higher SLR had a higher prevalence of proteinuria, and individuals with eGFR < 60/min/1.73m^2^ had a higher prevalence (All *P* < 0.05).

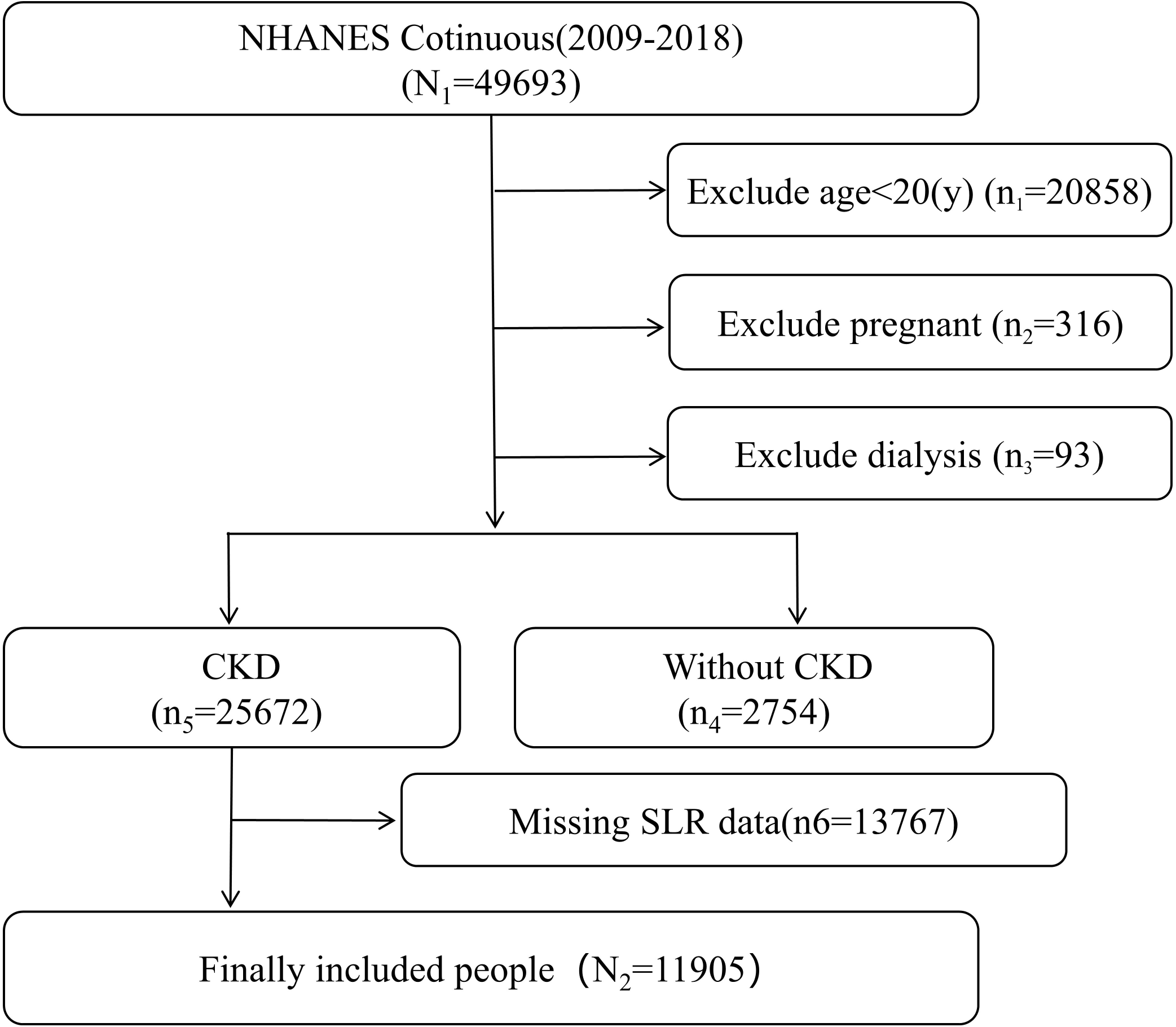

### 3.2 Association of SLR with the risk of CKD

Patients were categorized into Tertiles 1 (T1), Tertiles 2 (T2), and Tertiles 3 (T3) based on SLR levels, representing lower SLR group, medium SLR group, and higher SLR group. RCS plot results showed a non-linear relationship between SLR and CKD risk, with increasing SLR levels associated with increased CKD risk in the general population (nknot=5, Non-line *P* value <0.01) **(Figure 2)**.

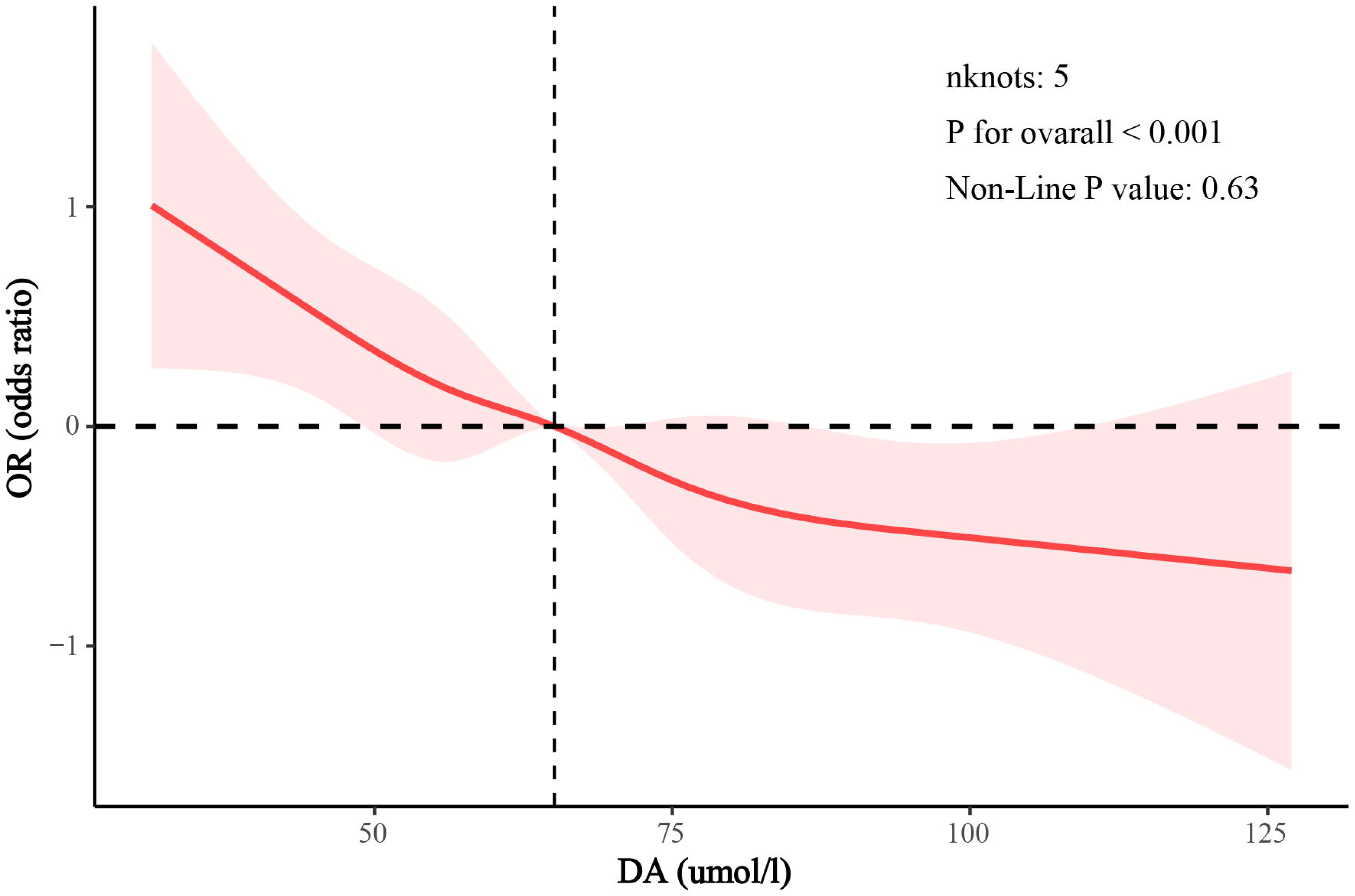

Univariate logistic regression showed that compared to the T1 group, the risk of CKD increased by 59% in the T2 group (OR = 1.59; 95% CI, 1.30-1.95; *P* < 0.001), and by 146% in the T3 group (OR = 2.46; 95% CI, 1.99-3.04; *P* < 0.001) **(Figure 3; Supplementary-table 2, 3)**. After adjusting for baseline age, gender, race, and BMI, Model 1 showed that individuals in the T2 group had a 35% higher risk of CKD compared to the T1 group (OR = 1.35; 95% CI, 1.07-1.70; *P* = 0.01), and individuals in the T3 group had a 93% higher risk (OR = 1.93; CI, 1.48-2.51; *P* < 0.001) **(Figure 3; Supplementary-table 2, 3)**. Model 2 included adjustments for the covariates in Model 1 as well as alcohol use(‘yes’ or ‘no’) and smoking(‘yes’ or ‘no’), showing that individuals in the T3 group had an 84% higher risk of CKD compared to the T1 group (OR = 1.84; 95% CI, 1.39-2.44; *P* < 0.001) **(Figure 3; Supplementary-table 2, 3)**. Model 3 further adjusted for the covariates in Model 2 as well as anemia (‘yes’ or ‘no’), hyperlipidemia (‘yes’ or ‘no’), hypertension (‘yes’ or ‘no’), diabetes mellitus (DM) (‘yes’ or ‘no’), and renin-angiotensin-aldosterone system inhibitors (RAASi) ("yes" or "no"), revealing that individuals in the T3 group had a 54% higher risk of CKD compared to the T1 group (OR = 1.54; 95% CI, 1.16-2.06; *P* = 0.004). Additionally, multivariable Cox regression analysis showed that for per standard deviation (per-SD) increase in SLR, there was a 16% increase in CKD risk (OR = 1.16; 95% CI, 1.05-1.29; *P* = 0.005) **(Figure 3; Supplementary-table 2, 3)**.

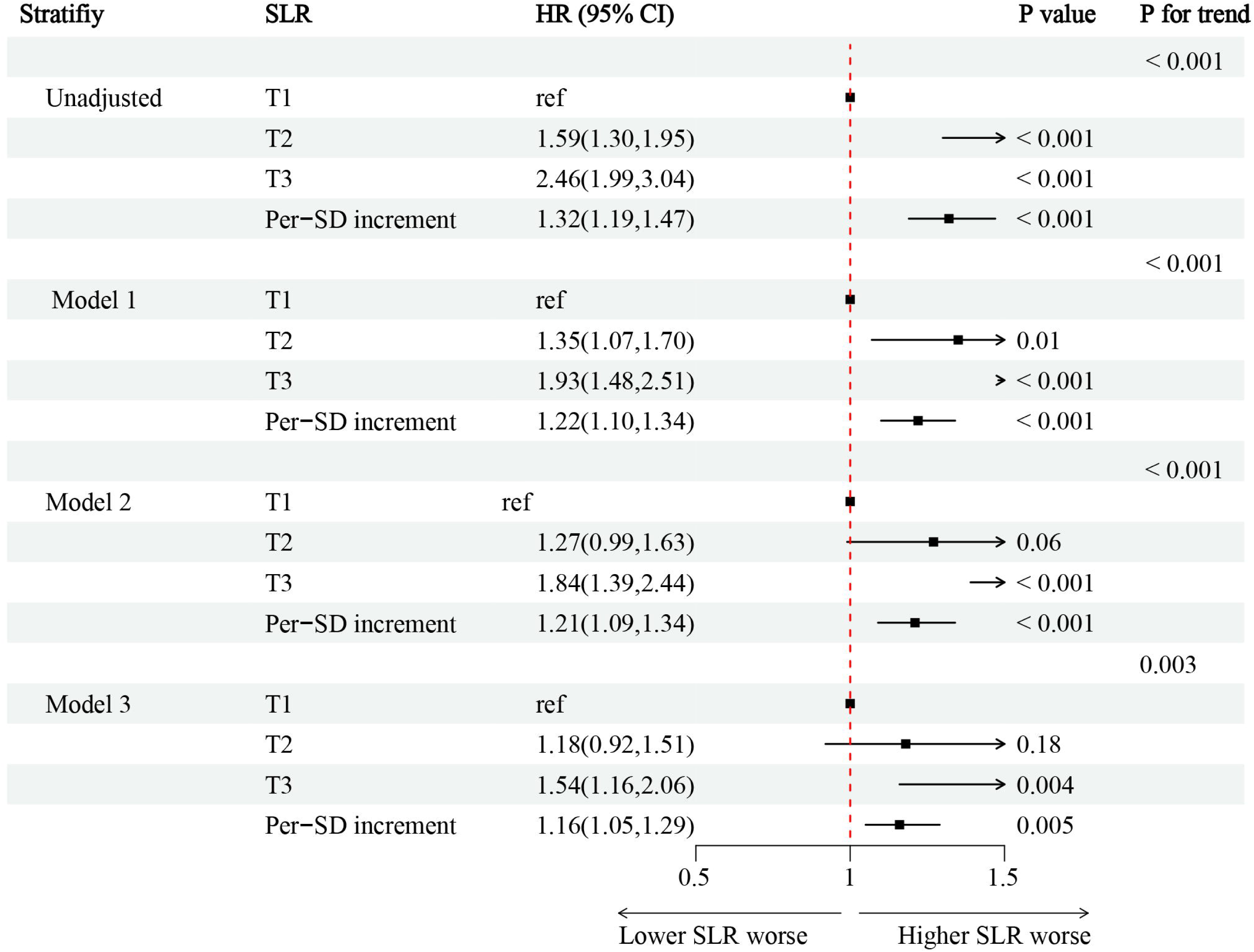

### 3.3 Stratified analysis

Stratified analysis indicated a significant gender-specific effect on the relationship between SLR and CKD risk (*P* for interaction < 0.05). However, there was no interaction with age (<60 years, ≥60 years), race, hypertension (’yes’ or ’no’), hyperlipidemia (’yes’ or ’no’), and diabetes (’yes’ or ’no’) (all *P* for interaction > 0.05) **(Table 2)**.

### 3.4 Mediation analysis

Mediation analysis is used to explore the mediating role of inflammation in the relationship between SLR and the risk of CKD in the general population. The association between SLR and CKD was mediated through SII (β = 0.01; 95% CI, 0.004-0.01; *P < 0.001*) **(Figure 4A)**. Furthermore, to ensure the accuracy of the results, we used SIRI to account for inflammation, and the results showed that the association between SLR and CKD can also be mediated through SIRI (β = 0.03; 95% CI, 0.02-0.03; *P < 0.001*) **(Figure 4B)**.

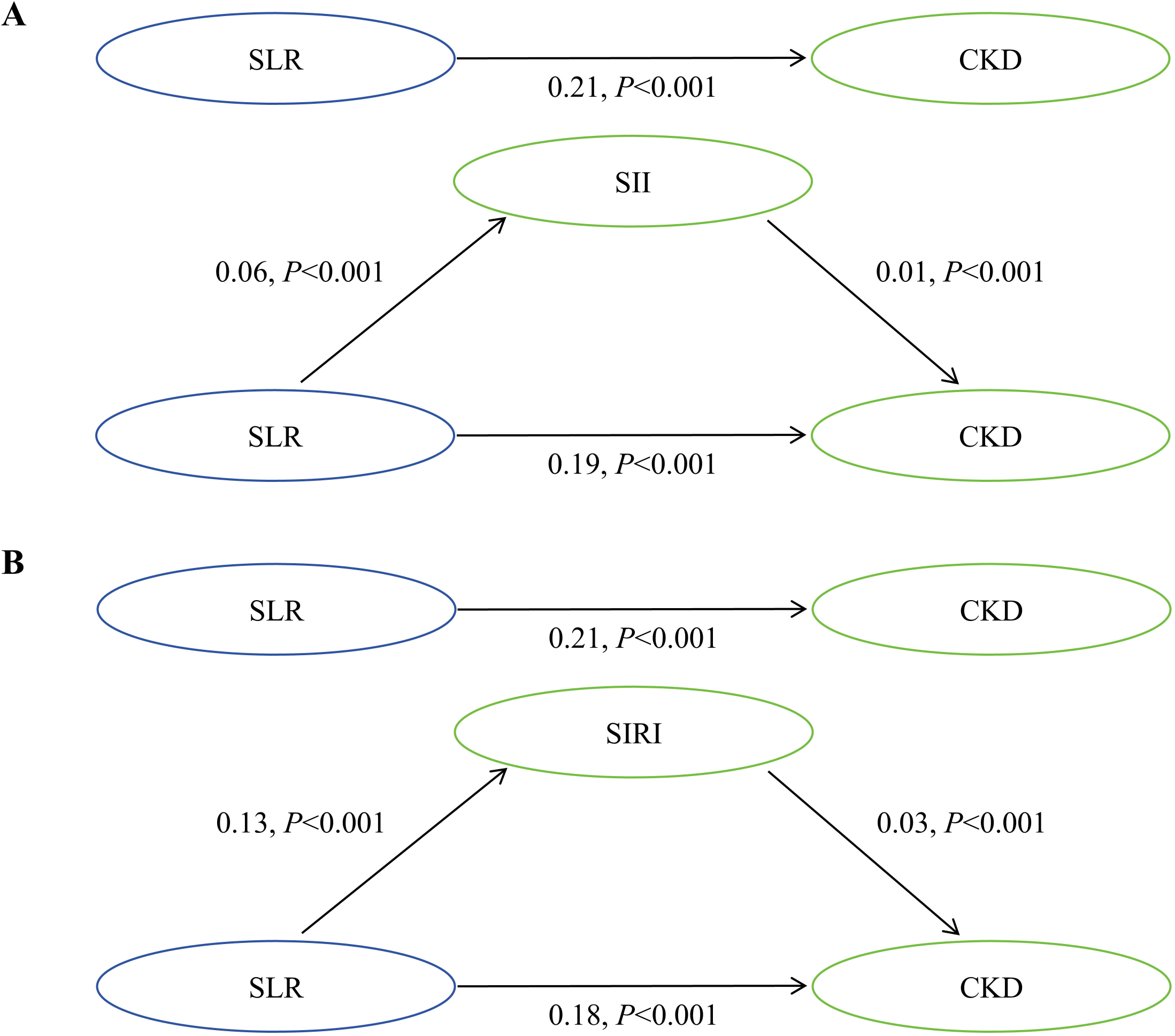

## 4. Discussion

Our study results showed a significant association between SLR and the risk of CKD in the general population of US, with higher SLR indicating an increased risk of CKD. This finding provides important insights for exploring the potential pathogenesis and risk assessment of CKD.

Recent studies have revealed the unique physiological role of sdLDL-C, which is closely associated with various diseases. sdLDL-C has been identified as the most significant risk factor for atherosclerotic cardiovascular disease (ASCVD) (22–24). The reason for this outcome may be that, unlike other LDL-C particles, small dense LDL cholesterol (sdLDL-C) exhibits reduced affinity for LDL receptors, resulting in prolonged plasma residence time. This allows sdLDL-C to penetrate the endothelium more easily, making it more susceptible to oxidative modification. Ultimately, it was taken up by macrophages to form foam cells. In addition, both in vivo and in vitro, sdLDL-C was more readily subject to glycosylation (8, 9, 23–26). Furthermore, sdLDL-C was strongly linked to diseases such as diabetes, carotid plaque formation, and MetS. Gerber PA et al. (27) found that the proportion of sdLDL-C particles could predict changes in insulin resistance in type 2 diabetes and was closely related to other determinants of metabolic status. Moreover, changes in sdLDL-C particle proportions could predict changes in carotid intimal medial thickness and insulin resistance (28). Conversely, the relationship between lbLDL-C particles and disease remains to be explored. Compared to sdLDL-C particles, lbLDL-C plays a weaker role in atherosclerosis.

However, in current study, SLR has been identified as an independent risk factor for MetS (15). This result may be attributed to the offsetting of individual subclass effects when LDL-C subclasses were combined, resulting in a lack of correlation between total LDL-C and MetS. Studies have also shown that in HIV-infected patients receiving long-term highly active antiretroviral therapy, SLR was a significant predictor of lipodystrophy, particularly effective in identifying a dysregulation of lipoprotein metabolism that typically increase cardiovascular disease risk (16). Additionally, SLR exhibited significant predictive ability for type 2 diabetes (T2D) (17).Our study demonstrates that SLR is an independent risk factor for CKD. However, the exact reasons for this finding are not yet fully understood. One possible explanation is that long-term circulating lipids metabolism imbalance can result in ectopic fat distribution in peripheral organs, especially in the heart, liver, and kidneys, with the kidneys being particularly susceptible to dyslipidemia (29).

Lipids accumulation induces tissue lipotoxicity, oxidative stress, fibrosis, and inflammation can result in glomerular and tubule-interstitial damage (29, 30). LDL-C may interfere with renal hemodynamics, affecting renal perfusion and filtration function. LDL-C has been associated with increased blood viscosity, thereby increasing vascular resistance (31, 32). Crowley, J. P. et al. (33) also found that LDL-C was an independent major lipoprotein affecting whole-blood viscosity, similar in effect to fibrinogen. This hemodynamic-mediated glomerular damage plays a crucial role in the pathogenesis of progress glomerulosclerosis (34–37). In addition, LDL-C might interact with other metabolic disturbances, forming a complex pathogenic network that further increases the risk of CKD. Studies have shown that ectopic lipid accumulation can exacerbate renal dysfunction, impair insulin signaling, mediate insulin resistance, leading to diabetic nephropathy (38, 39). Recently, SLR has been confirmed that correlated with MetS (15, 17), which has been shown to significantly increase the risk of CKD (40). Meanwhile, an increase in the SLR ratio indicates a rise in sdLDL-C (15), a subtype of cholesterol with stronger atherogenic properties, including glycosylation, oxidation, prolonged half-life period, reduced LDL receptor affinity, smaller particle size facilitating endothelial permeation (8, 9, 25, 41, 42), uptake by monocyte-derived macrophages to form foam cells, and smooth muscle cell proliferation resulting in atherosclerotic plaque formation. The atherosclerotic changes in renal vasculature undoubtedly have adverse effects on renal health. In addition, the SLR was particularly effective in identifying lipid metabolism disorders and predicting lipodystrophy(15). lipodystrophy was associated with an increased risk of CKD or kidney impairment in patients (43, 44).

Meanwhile, numerous studies indicate that sdLDL-C activates both innate and adaptive immune cells, leading to inflammatory responses through various mechanisms(45, 46).. Additionally, oxidatively modified LDL engages multiple macrophage receptors, triggering intracellular signaling cascades that promote inflammation. Due to its distinct physicochemical characteristics, sdLDL-C is more susceptible to oxidative modification, which may enhance its inflammatory potential(11). Our research further shows that inflammation plays a mediating role in the relationship between SLR and CKD, offering new insights into the mechanisms underlying CKD.

This study acknowledges several limitations. Firstly, as a cross-sectional analysis, while associations between the variables were identified, causality cannot be inferred. Secondly, additional influencing factors may exist that were not accounted for in the analysis, potentially affecting the results. Future research should employ more robust methodologies, larger sample sizes, and prospective designs to further validate and investigate this relationship. Lastly, the lack of direct measurements of sdLDL-C and lbLDL-C levels introduces the possibility of bias.

Despite these limitations, the findings suggest a novel biomarker for identifying individuals at high risk of CKD in the general population. Furthermore, inflammation plays a mediating role in the relationship between different subtypes of LDL-C and the risk of CKD, which also provides new insights into the mechanisms by which lipid metabolism abnormalities lead to the onset and progression of CKD.

## Data Availability

All data generated in this study is available at the reasonable request of the authors, all data generated in this study is included in the manuscript, and all data generated is available online by accessing the NHANES database.

https://wwwn.cdc.gov/nchs/nhanes/Default.aspx

## Funding

The Science and technology fund of Chengdu Medical College (CYZYB22-02).

The research fund of Sichuan Medical and Health Care Promotion institute (KY2022QN0309).

The Sichuan Provincial Medical Association Youth Innovation Project (Q23021).

Project of Sichuan Provincial Science and Technology Department (2023-4-763).

## Author Contributions

Conception and design of the study: Xiang Xiao, Tingyu Wu, Rong Ma, Yujun Tang, Zhiqiang

Duan. Acquisition and analysis of data: Xiang Xiao, Yun Sun, Rong Ma, Tingyu Wu; Drafting the manuscript or figures: Xiang Xiao, Yujun Tang, Rong Ma, Wan Li, Pin Zhou, Zhuoxing Li, Donglin Liu, Haiyan Yu, Guifei Deng, Qiyuan Liang.

## Data Availability

Some or all datasets generated during and/or analyzed during the current study are not publicly available but are available from the corresponding author on reasonable request.

## Acknowledgments

The authors wish to thank all the participants of this study for their important contributions.

## Ethical Approval

The study protocol conformed to the ethical standards of the 1964 Declaration of Helsinki and its subsequent amendments, with approval from the National Committee for Ethical Review of Health Statistics Research and signed informed consent from all participants.

## Declarations

The authors declare that they have no known competing financial interests or personal relationships that could have appeared to influence the work reported in this paper.

## Notes

### Competing Interest Statement

The authors have declared no competing interest.

