## Supplementary material for "Relationship Between Ratio of Low-Density Lipoprotein Cholesterol subtypes and Risk of Chronic Kidney Disease: The mediating role of inflammation": Table 1: Table_SLR.docx

**Table 1. Baseline clinical features of enrolled individuals**

| Variable | Total (N=11905) | T1  (≤0.37)  (n_1_=3969) | T2  (0.37-0.51)  (n_2_=3969) | T3  (＞0.37)  (n_3_=3967) | *P*-value |
| --- | --- | --- | --- | --- | --- |
| SLR | 0.47(0.00) | 0.30(0.00) | 0.44(0.00) | 0.67(0.01) | < 0.001 |
| SII | 507.81(4.59) | 481.63(7.40) | 516.58(6.57) | 525.77(6.43) | < 0.001 |
| SIRI | 1.19(0.01) | 1.07(0.02) | 1.21(0.02) | 1.29(0.02) | < 0.001 |
| Age (years) | 48.29(0.29) | 45.11(0.43) | 48.42(0.38) | 51.40(0.40) | < 0.001 |
| Gender(%) |  |  |  |  | < 0.001 |
| Female | 51.48(0.01) | 56.15(1.03) | 53.16(0.95) | 44.99(1.00) |  |
| Male | 48.52(0.01) | 43.85(1.03) | 46.84(0.95) | 55.01(1.00) |  |
| Race (%)2 |  |  |  |  | < 0.001 |
| Mexican American | 8.62(0.01) | 6.49(0.67) | 9.44(1.06) | 9.96(0.93) |  |
| Non-Hispanic Black | 10.43(0.01) | 16.03(1.07) | 9.49(0.73) | 5.65(0.56) |  |
| Non-Hispanic White | 66.27(0.03) | 63.07(1.68) | 66.69(1.68) | 69.10(1.53) |  |
| Other Hispanic | 6.27(0.01) | 5.97(0.69) | 6.20(0.62) | 6.65(0.71) |  |
| Other Race - Including Multi-Racial | 8.42(0.01) | 8.44(0.71) | 8.18(0.61) | 8.64(0.69) |  |
| BMI (%) |  |  |  |  | < 0.001 |
| Underweight | 1.62(0.00) | 2.42(0.34) | 1.51(0.25) | 0.95(0.22) |  |
| Normal weight | 27.89(0.01) | 41.11(1.32) | 27.05(1.20) | 15.82(0.76) |  |
| Overweight | 32.16(0.01) | 31.80(1.02) | 33.88(1.03) | 31.46(0.93) |  |
| Obesity | 37.63(0.01) | 24.67(1.03) | 37.56(1.05) | 51.77(1.01) |  |
| Alcohol use (%) |  |  |  |  | 0.33 |
| No | 9.54(0.01) | 10.70(0.80) | 11.30(0.69) | 10.01(0.79) |  |
| Yes | 79.90(0.02) | 89.30(0.80) | 88.70(0.69) | 89.99(0.79) |  |
| Smoke (%) |  |  |  |  | < 0.001 |
| No | 55.70(0.02) | 62.49(1.14) | 55.52(1.21) | 48.99(1.21) |  |
| Yes | 44.28(0.02) | 37.51(1.14) | 44.48(1.21) | 51.01(1.21) |  |
| Serum albumin (g/L) | 42.34(0.07) | 42.69(0.09) | 42.22(0.09) | 42.12(0.10) | < 0.001 |
| ACR (mg/g) | 31.40(1.99) | 16.38(1.14) | 29.56(3.54) | 48.74(5.14) | < 0.001 |
| ACR (mg/g) |  |  |  |  | < 0.001 |
| <30 | 90.19(0.03) | 93.22(0.53) | 90.85(0.69) | 87.12(0.72) |  |
| (30-300) | 8.13(0.00) | 6.08(0.49) | 7.79(0.64) | 10.65(0.66) |  |
| ≥300 | 1.42(0.00) | 0.70(0.14) | 1.36(0.21) | 2.23(0.29) |  |
| Anemia (%) |  |  |  |  | 0.39 |
| No | 93.28(0.03) | 93.05(0.50) | 93.57(0.54) | 93.99(0.50) |  |
| Yes | 6.45(0.00) | 6.95(0.50) | 6.43(0.54) | 6.01(0.50) |  |
| Hypertension (%) |  |  |  |  | 0.18 |
| No | 61.29(0.02) | 59.99(1.27) | 62.88(1.10) | 60.98(1.24) |  |
| Yes | 38.71(0.01) | 40.01(1.27) | 37.12(1.10) | 39.02(1.24) |  |
| DM |  |  |  |  | < 0.001 |
| NO | 83.41(0.02) | 93.19(0.49) | 85.34(0.71) | 71.48(1.04) |  |
| Yes | 16.59(0.01) | 6.81(0.49) | 14.66(0.71) | 28.52(1.04) |  |
| Hyperlipidemia (%) |  |  |  |  | < 0.001 |
| No | 30.00(0.01) | 47.43(1.08) | 31.70(1.07) | 10.47(0.78) |  |
| Yes | 70.00(0.02) | 52.57(1.08) | 68.30(1.07) | 89.53(0.78) |  |
| e-GFR (ml/min/1.73m^2^) |  |  |  |  | < 0.001 |
| >60 | 93.40(0.03) | 96.68(0.35) | 93.57(0.53) | 89.89(0.58) |  |
| <60 | 6.60(0.00) | 3.32(0.35) | 6.43(0.53) | 10.11(0.58) |  |
| RAASi use (%) |  |  |  |  | < 0.001 |
| No | 80.59(0.02) | 88.22(0.89) | 81.68(0.88) | 71.89(0.98) |  |
| Yes | 19.32(0.01) | 11.78(0.89) | 18.32(0.88) | 28.11(0.98) |  |

SLR, Small Dense Low-Density Lipoprotein Cholesterol/Large Buoyant Low-Density Lipoprotein Cholesterol Ratio; SII, Systemic Immune-Inflammation Index; SIRI, Systemic Inflammation Response Index; BMI, Body Mass Index; ACR, albumin-to-creatinine ratio; DM, diabetes mellitus; e-GFR, estimated glomerular filtration rate; RAASi, renin-angiotensin system inhibitors.

**Table 2. Stratified analysis of sdLDLand the risk of CKD in individuals**

| Variable | OR（95% CI） | | | | *P* for interaction |
| --- | --- | --- | --- | --- | --- |
|  | T1 | T2 | T3 | *P* for trend |  |
| Age (years) |  |  |  |  | 0.94 |
| <60 | ref | 1.31(0.95,1.79) | 1.93(1.41,2.64) | <0.001 |  |
| ≥60 | ref | 1.35(0.96,1.91) | 1.99(1.41,2.80) | <0.001 |  |
| Gender |  |  |  |  | 0.02 |
| Male | ref | 0.89(0.64,1.24) | 1.61(1.13,2.28) | 0.003 |  |
| Female | ref | 1.57(1.21,2.02) | 1.91(1.41,2.57) | <0.001 |  |
| Race |  |  |  |  | 0.28 |
| Non-Hispanic White | ref | 1.25(0.75,2.09) | 1.61(0.99,2.61) | 0.04 |  |
| Other Race - Including Multi-Racial | ref | 1.34(0.99,1.82) | 1.93(1.37,2.73) | <0.001 |  |
| Non-Hispanic Black | ref | 1.17(0.80,1.71) | 2.05(1.46,2.87) | <0.001 |  |
| Mexican American | ref | 1.39(0.66,2.95) | 1.42(0.69,2.93) | 0.33 |  |
| Other Hispanic | ref | 0.62(0.31,1.25) | 0.88(0.47,1.64) | 0.79 |  |
| Hyperlipidemia |  |  |  |  | 0.25 |
| No | ref | 1.22(0.94,1.58) | 1.64(1.23,2.20) | <0.001 |  |
| Yes | ref | 1.33(0.90,1.96) | 2.95(1.66,5.24) | 0.001 |  |
| Hypertension |  |  |  |  | 0.81 |
| No | ref | 1.08(0.81,1.44) | 1.48(1.01,2.15) | 0.03 |  |
| Yes | ref | 1.34(0.95,1.89) | 1.83(1.31,2.57) | <0.001 |  |
| DM |  |  |  |  | 0.47 |
| N0 | ref | 1.11(0.84,1.47) | 1.55(1.16,2.08) | 0.004 |  |
| Yes | ref | 1.28(0.91,1.80) | 1.35(0.95,1.91) | 0.19 |  |

OR, odds ratio; CI, Confidence interval; DM, diabetes mellitus.

**Figure 1.** Enrollment of flowchart.

**Figure 2.** Association between SLR and the risk of CKD in individuals based on restricted cubic spline plot.

**Figure 3.** Associations between SLR level and the risk of CKD in individuals . **Model 1** adjusted for baseline age, gender, race, BMI; **Model 2** adjusted for covariates in model 1 plus , smoke (‘yes’ or ‘no’), alcohol use (‘yes’ or ‘no’); **Model 3** adjusted for covariates in model 2 plus anemia (‘yes’ or ‘no’), hyperlipidemia (‘yes’ or ‘no’), hypertension (‘yes’ or ‘no’)，DM (‘yes’ or ‘no’), RAASi use (‘yes’ or ‘no’). CKD, chronic kidney disease; OR, Odd Ratio; CI, confidence interval; BMI, Body Mass Index; DM, diabetes mellitus; RAASi, renin-angiotensin system inhibitors.

**Figure 4.** Inflammatory markers mediated the relationship between SLR and CKD: A. SII); B. SIRI. SII, Systemic Immune-Inflammation Index; SIRI, Systemic Inflammation Response Index.
