## Supplementary table 1 for "Relationship Between Ratio of Low-Density Lipoprotein Cholesterol subtypes and Risk of Chronic Kidney Disease: The mediating role of inflammation": Supplementary_SLR.docx

**Supplementary -table 1. Definitions/criteria of some diagnoses**

| Variables | Definitions/criteria |
| --- | --- |
| Smoker | Smoking more than 100 cigarettes in previous and now. |
| Alcohol user ^1^ | ≥2 drinks per day for females, ≥3 drinks per day for males, or binge drinking ≥2 days per month.  Binge drinking (≥4 drinks on the same occasion for females, ≥5 drinks on the same occasion for males) on 5 or more days per month. |
| Hypertension ^2^ | (1) Self-reported hypertension diagnosis, (2) Use of anti-hypertensive medication, (3) Average systolic blood pressure (SBP) > 140 mmHg, (4) Average diastolic blood pressure (DBP) > 90 mmHg, meet any of the above conditions. |
| Anemia ^3^ | ≥120g/L for women (15 years of age and above), ≥130g/L for men (15 years of age and above). |
| Hyperlipidemia | (1) Triglyceridemia ≥ 150 mg/dl; (2) Hypercholesterolemia: a) total cholesterol ≥ 200 mg/dl, b) low-density lipoprotein ≥ 130 mg/dl), c). high-density lipoprotein (< 40 mg/dl, male; < 50 mg/dl, female), meet any of the above conditions; (3) Use of lipid-lowering drugs; meet any of the above conditions. |
| Diabetes mellitus | 1) a doctor has told you that you have diabetes, 2) HbA1c (%) > 6.5, 3) fasting blood glucose (mmol/l) ≥ 7.0, 4) random blood glucose (mmol/l) ≥ 11.1, and 5) two-hour oral glucose tolerance test (OGTT) blood glucose (mmol/l) >= 11.1 ^4^. |

**Supplementary -table 2. Logistic-regression analysis of risk factors for CKD in individuals**

| Variables | Unadjusted | | Model 3 | |
| --- | --- | --- | --- | --- |
|  | OR (95% CI) | *P*-value | OR (95% CI) | *P*-value |
| SLR |  |  |  |  |
| T1 | ref |  | ref |  |
| T2 | 1.59(1.30,1.95) | <0.001 | 1.18(0.92,1.51) | 0.18 |
| T3 | 2.46(1.99,3.04) | <0.001 | 1.54(1.16,2.06) | 0.004 |
| Age | 1.06(1.05,1.07) | <0.001 | 1.04(1.02, 1.06) | 0.01 |
| Gender |  |  |  |  |
| Female | ref |  | ref |  |
| Male | 0.81(0.65,1.02) | 0.07 | 0.82(0.51, 1.33) | 0.23 |
| Race |  |  |  |  |
| Mexican American | ref |  | ref |  |
| Non-Hispanic Black | 2.01(1.19,3.41) | 0.12 | 0.92(0.30, 2.77) | 0.76 |
| Non-Hispanic White | 3.03(1.89,4.85) | 0.56 | 0.59(0.21, 1.65) | 0.16 |
| Other Hispanic | 1.51(0.75,3.01) | 0.99 | 0.71(0.21, 2.42) | 0.35 |
| Other Race - Including Multi-Racial | 1.68(0.82,3.43) | 0.85 | 0.66(0.24, 1.86) | 0.23 |
| BMI | 0.95(0.76,1.19) | 0.66 | 0.92(0.60, 1.40) | 0.48 |
| Alcohol use |  |  |  |  |
| No | ref |  | ref |  |
| Yes | 0.64(0.44,0.94) | 0.02 | 0.83(0.43, 1.58) | 0.34 |
| Smoke |  |  |  |  |
| No | ref |  | ref |  |
| Yes | 1.34(0.97,1.84) | 0.07 | 1.23(0.58, 2.61) | 0.35 |
| Anemia |  |  |  |  |
| No | ref |  | ref |  |
| Yes | 3.48(2.71,4.45) | <0.001 | 2.34(1.25, 4.39) | 0.03 |
| Hyperlipidemia |  |  |  |  |
| No | ref |  | ref |  |
| Yes | 2.60(1.76,3.86) | <0.001 | 1.38(0.46, 4.16) | 0.34 |
| Hypertension |  |  |  |  |
| No | ref |  | ref |  |
| Yes | 4.17(3.13,5.55) | <0.001 | 1.82(1.01, 3.26) | 0.05 |
| DM |  |  |  |  |
| No | ref |  | ref |  |
| Yes | 5.09(4.14,6.24) | <0.001 | 2.75(1.64, 4.60) | 0.01 |

### OR, odds ratio; CI, Confidence interval; BMI, body mass index; RAASi, renin-angiotensin system inhibitors; CKD, chronic kidney disease; DM, Diabetes mellitus.

**Supplementary -table 3. Multifactor logistic regression analysis of risk factors for the risk of** **CKD in individuals**

| Variables | Unadjusted | | Model 1 | | Model 2 | | Model 3 | |
| --- | --- | --- | --- | --- | --- | --- | --- | --- |
|  | 95% CI | P-value | 95% CI | P-value | 95% CI | P-value | 95% CI | P-value |
| T1 | ref |  | ref |  | ref |  | ref |  |
| T2 | 1.59(1.30,1.95) | <0.001 | 1.35(1.07,1.70) | 0.01 | 1.27(0.99,1.63) | 0.06 | 1.18(0.92,1.51) | 0.18 |
| T3 | 2.46(1.99,3.04) | <0.001 | 1.93(1.48,2.51) | <0.001 | 1.84(1.39,2.44) | <0.001 | 1.54(1.16,2.06) | 0.004 |
| P for trend |  | <0.001 |  | <0.001 |  | <0.001 |  | 0.003 |
| Per-SD increment of SLR | 1.32(1.19,1.47) | <0.001 | 1.22(1.10,1.34) | <0.001 | 1.21(1.09,1.34) | <0.001 | 1.16(1.05,1.29) | 0.005 |

**Model 1*^a^*** adjusted for baseline age, gender, race, BMI; **Model 2*^b^*** adjusted for covariates in model 1 plus smoke (‘yes’ or ‘no’), alcohol use (‘yes’ or ‘no’). **Model 3*^c^*** adjusted for covariates in model 2 plus anemia (‘yes’ or ‘no’), hyperlipidemia (‘yes’ or ‘no’), hypertension (‘yes’ or ‘no’)，DM (‘yes’ or ‘no’), RAASi use (‘yes’ or ‘no’). OR, odds ratio; CI, Confidence interval; BMI, Body Mass Index; RAASi, renin-angiotensin system inhibitors; CKD, chronic kidney disease; DM, diabetes mellitus.
